# Exploring Undergraduates’ Knowledge, Attitude, and Perception of Infertility in Osun State University: A mixed method study

**DOI:** 10.64898/2026.03.30.26349746

**Authors:** Kehinde Awodele, Sunday Charles Adeyemo, Adesegun Tajudeen Waliu, Adeniyi Olaonipekun Fasanu, Busayo Temilola Akinbowale, Victoria Adenike Adeniyi, Roseline Folami, Oladayo Damilola Akinwale, Joshua Falade, Eniola Dorcas Olabode

**Affiliations:** Department of Obstetrics and Gynecology, Osun State University, Osogbo, Nigeria; Health and Biomedical Sciences, Institut Superiuer De Santer, Niamey, Niger; Department of Public Health, Osun State University, Osogbo, Nigeria; Faculty of Nursing Sciences, Osun State University, Osogbo, Nigeria; Department of Mental Health, University of Medical Sciences Ondo, Nigeria

**Keywords:** Undergraduates, Male Infertility, Knowledge, Attitude, Perception, Osun State University, Nigeria

## Abstract

**Background:** Conventionally, infertility has been regarded as primarily a female issue, leading to misconceptions, stigma, and underrepresentation of male infertility in healthcare discussions. This study assessed the knowledge, attitude and perception of Undergraduates towards male infertility in Osun State University.

**Methods:** A descriptive cross-sectional design was employed to select 300 undergraduates via multistage sampling. Qualitative data were collected using a focus group discussion guide covering the knowledge, attitude and perception, while quantitative data were collected using a self-administered questionnaire covering socio-demographic characteristics, knowledge, attitude and perception towards male infertility. Qualitative analysis was performed using NVivo software, while IBM SPSS Statistics version 27 was used for the quantitative analysis, with thematic analysis and chi-square tests to determine the association between variables (significance at p < 0.05).

**Results:** Respondents were predominantly females (64.0%) with a mean age of 20.99 ± 2.31 years. Overall knowledge was low (47.7%), while more than half had a negative attitude (52.3%). Significant predictors of attitude include faculty (0.049), level (p=0.031), and formal education on male infertility (p=0.007).

**Conclusion:** Students demonstrated a poor understanding of male infertility, and their attitudes remain influenced by cultural norms surrounding marriage, masculinity, and gender roles. Hence, the need to foster open dialogues, promote gender-inclusive narratives, and strengthen healthcare support systems.

## INTRODUCTION

Infertility is defined as the inability of a couple to conceive after one year of consistent, unprotected sexual intercourse [1]. Traditionally, it has been viewed as primarily a female issue, leading to misconceptions, stigma, and the underrepresentation of male infertility in healthcare discussions [2]. This misconception has resulted in a lack of awareness and reluctance among men to seek medical assistance, exacerbating the social and psychological impact on affected individuals and families [3]. Recent qualitative evidence from fertility centers in Osogbo, Nigeria, confirms that stigma often leads to male infertility being perceived as a female issue, resulting in delayed diagnosis and treatment [4].

Worldwide, 10–15% of couples experience infertility, with male factors contributing to approximately 50% of cases [1]. Several publications report systematic declines in sperm counts over recent decades, highlighting the increasing influence of the male factor on the global prevalence of infertility [5]. In Nigeria, recent studies have documented a high burden of male infertility, with 84.3% of men attending fertility clinics in Osogbo having at least one abnormality in their seminal fluid analysis. Common abnormalities reported include asthenospermia (66.9%), teratospermia (63.9%), and oligospermia (21.0%) [6].

In Sub-Saharan Africa, male infertility affects an estimated 20–35% of couples seeking fertility treatment, a rate significantly higher than in developed countries [7]. Contributing factors include genetic abnormalities, hormonal imbalances, infections, and environmental exposures [8]. In Nigeria, cultural beliefs and societal norms further reinforce gender biases in reproductive health, often excluding men from infertility discussions [9]. The psychosocial consequences of male infertility are profound, with affected men experiencing psychological distress, emotional turmoil, and social marginalization [4].

University students represent a population on the cusp of making reproductive decisions. However, research suggests that infertility education is often inadequate, even in medical curricula [10]. Despite the increasing burden of male infertility in Nigeria, as evidenced by high prevalence rates of abnormal seminal fluid parameters [6, 11] and significant psychosocial consequences for affected men [4], limited empirical research exists on the awareness, knowledge, and perceptions of this condition among Nigerian undergraduates. This study aims to fill that gap by assessing how well students understand male infertility, identifying factors influencing their perceptions, and highlighting areas for educational improvement. Understanding these perceptions is crucial for fostering a more inclusive approach to infertility management and reducing associated stigma, particularly given that many of these students will soon be making reproductive decisions that could be informed by their understanding of male factor infertility.

## METHODOLOGY

### Study Setting and Design

This cross-sectional study was conducted at Osun State University (UNIOSUN), a multi-campus institution in Nigeria. The university has six campuses, with the main campus in Osogbo housing the College of Health Sciences [12]. A mixed-methods design, combining quantitative and qualitative approaches, was employed.

### Study Population and Sampling

The study population comprised all registered undergraduates at UNIOSUN. Students who were unwilling to participate or were too ill were excluded. A multistage sampling method was used. The sample size was calculated using Cochran’s formula, based on a prevalence of 73% for a positive attitude towards male infertility from a previous study [13], with a 95% confidence interval and a 10% non-response rate. The final sample size was 333. Simple random sampling (balloting) was used to select final participants.

### Data Collection

Data was collected from June to July 2025. Quantitative data were collected using a structured, self-administered questionnaire. The questionnaire included sections on socio-demographics, knowledge (4 questions, maximum score 10), attitude (4 questions, maximum score 4), and perception of male infertility. A score of ≥8 was classified as “good knowledge,” and a score of ≥2 as a “positive attitude” [10]. Qualitative data were gathered through four focus group discussions (FGDs) with 8–10 participants each, using a semi-structured guide. All FGDs were audio-recorded and transcribed verbatim.

### Data Analysis

Quantitative data were analyzed using IBM SPSS Statistics version 27. Descriptive statistics (frequencies, percentages, means) were calculated. Chi-square tests and binary logistic regression were used to determine associations between variables, with statistical significance set at p < 0.05. Qualitative data were analyzed using thematic analysis with NVivo software. Codes were generated inductively and grouped into themes related to knowledge, attitude, and perception.

### Ethical Considerations

Ethical approval was obtained from Osun State University (UNIOSUN) Research Ethics Committee (UTH/REC/2025/1285). Written informed consent was obtained from all participants before data collection. Confidentiality was ensured by anonymizing all data. The ethical principles and guidelines set out by the Declaration of Helsinki, the Belmont Report, and other relevant documents were followed during the conduct of the study.

## RESULTS

### Quantitative Study

A total of 333 questionnaires were administered, with 320 returned, giving a response rate of 90%. After excluding 20 questionnaires due to missing data, 300 were retained for analysis.

### Sociodemographic characteristics

The respondents were between the ages of 16-30, with a mean of 20.99 ± 2.31 years. The majority of the respondents (193, 64.0%) were females, the majority (290, 96.7%) were in Health sciences, more than half (173, 57.7%) were in 400 level, almost all of the respondents (289, 96.3%) were single, and a large proportion 198 (66.0%) had received formal education on male infertility. (Table 1)

**Table 1:** Sociodemographic Characteristics of the Respondents (N=300)

| Variable | Frequency | Percentage |
| --- | --- | --- |
| **Gender** |  |  |
| Male | 107 | 35.7 |
| Female | 193 | 64.3 |
| **Age** |  |  |
| 16-20 | 144 | 48 |
| 21-25 | 145 | 48.3 |
| 26-30 | 11 | 3.7 |
| **Faculty** |  |  |
| Health Sciences | 290 | 96.7 |
| Social Sciences | 3 | 1.0 |
| Others | 7 | 2.3 |
| **Level of study** |  |  |
| 100 | 4 | 1.3 |
| 200 | 57 | 19.0 |
| 300 | 61 | 20.3 |
| 400 | 173 | 57.7 |
| 500 | 5 | 1.7 |
| **Marital status** |  |  |
| Single | 289 | 96.3 |
| Married | 11 | 3.6 |
| **Ever received formal education on male infertility?** |  |  |
| Yes | 198 | 66.0 |
| No | 102 | 34.0 |

### Respondents’ perception of male infertility

The majority 258 (86.0%) believe it can be treated, a large proportion 221 (73.7%) think it is not widely discussed in Nigeria, and major barriers to seeking treatment includes; sigma and cultural belief (258, 86.0%), lack of awareness (259, 86.3%), fear of being judged (243, 81.0%). (Table 2)

**Table 2:**
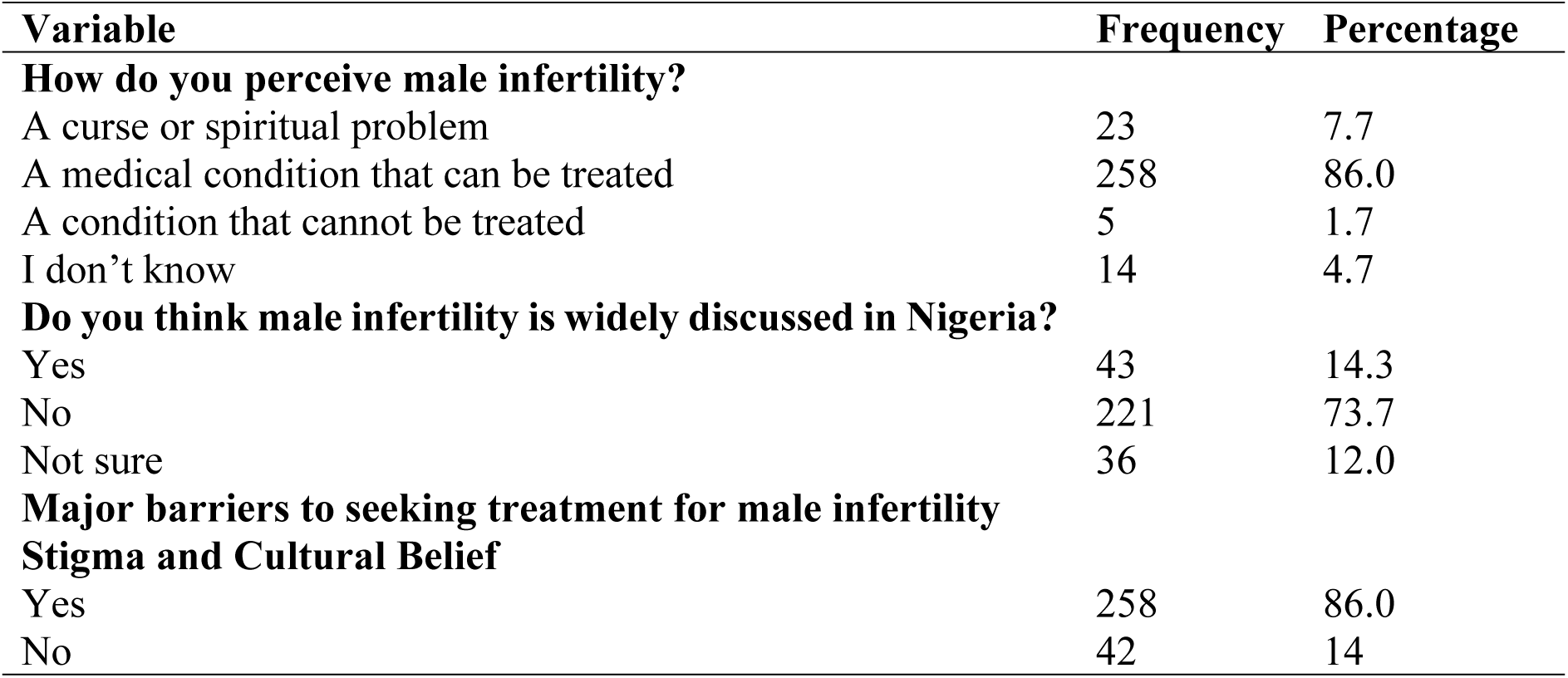
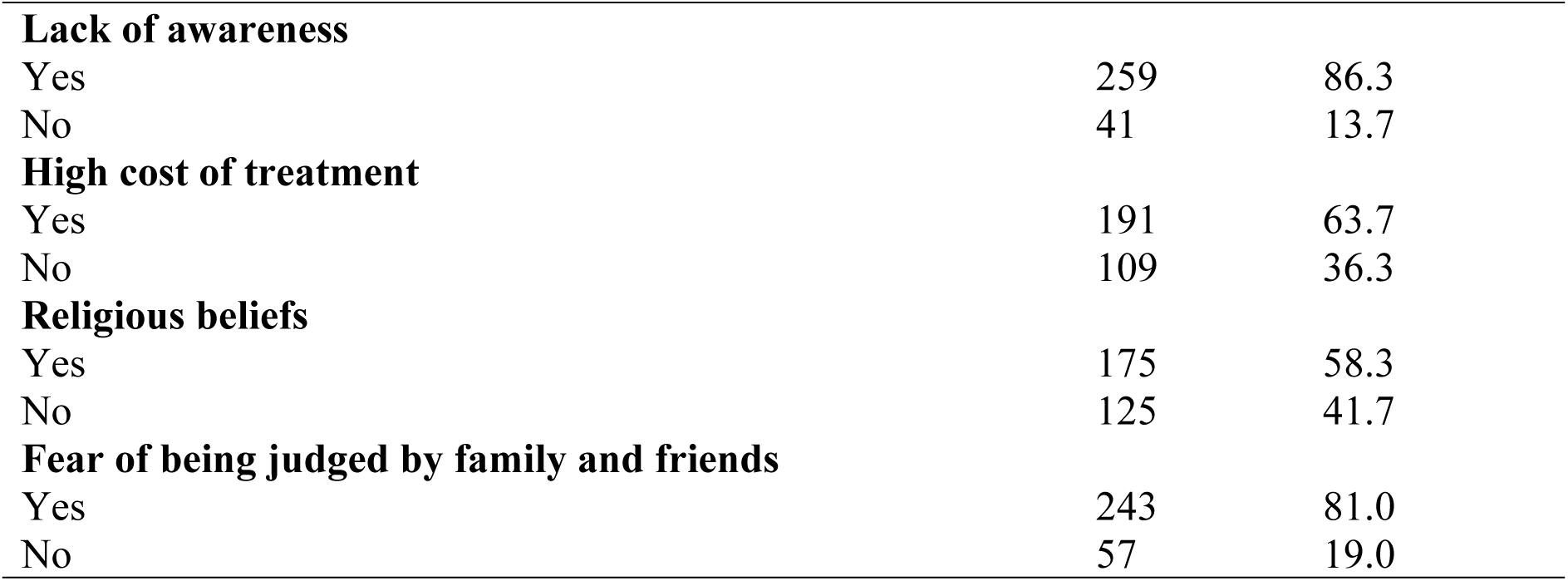
Respondents’ Perception of Male Infertility (N=300)

Association between respondents’ knowledge and attitude levels regarding male infertility and their sociodemographic characteristics. Although not statistically significant, slightly more than half of male respondents (50.5%) demonstrated good knowledge of male infertility, compared to less than half of female respondents (46.1%). Notably, respondents who had received formal education on male infertility were more likely to demonstrate good knowledge (51.0%) compared to those who had not (41.2%), though this difference did not reach statistical significance (Table 3).

**Table 3:**
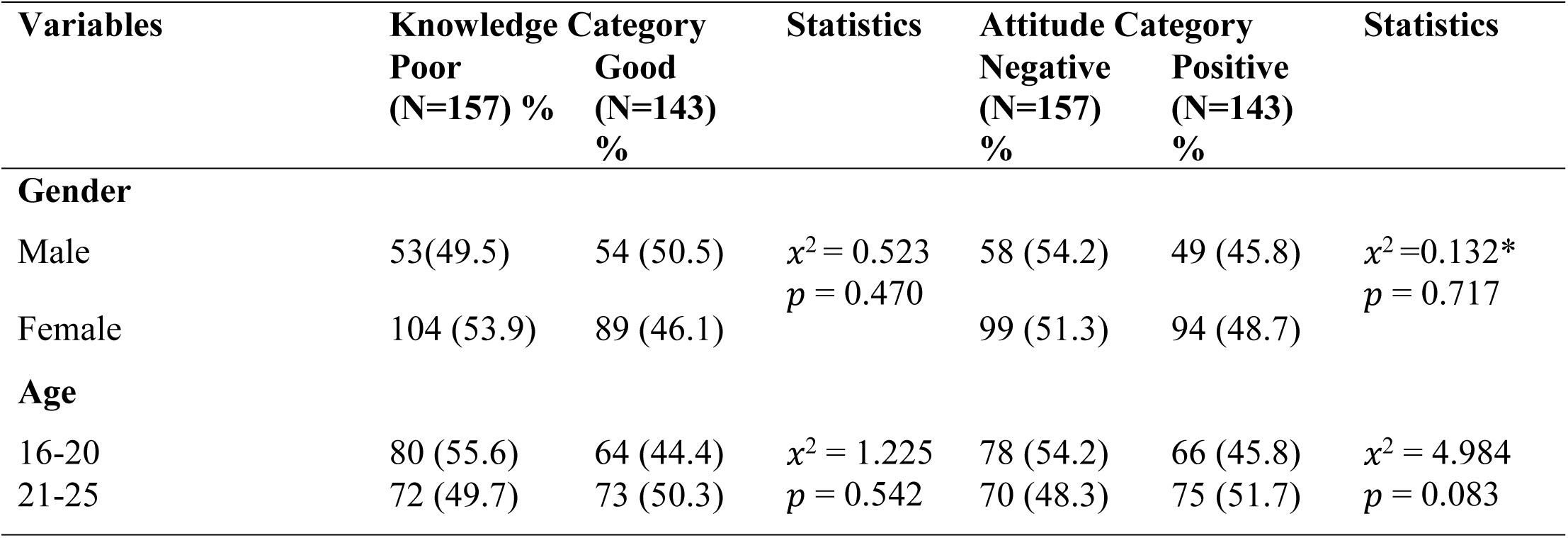
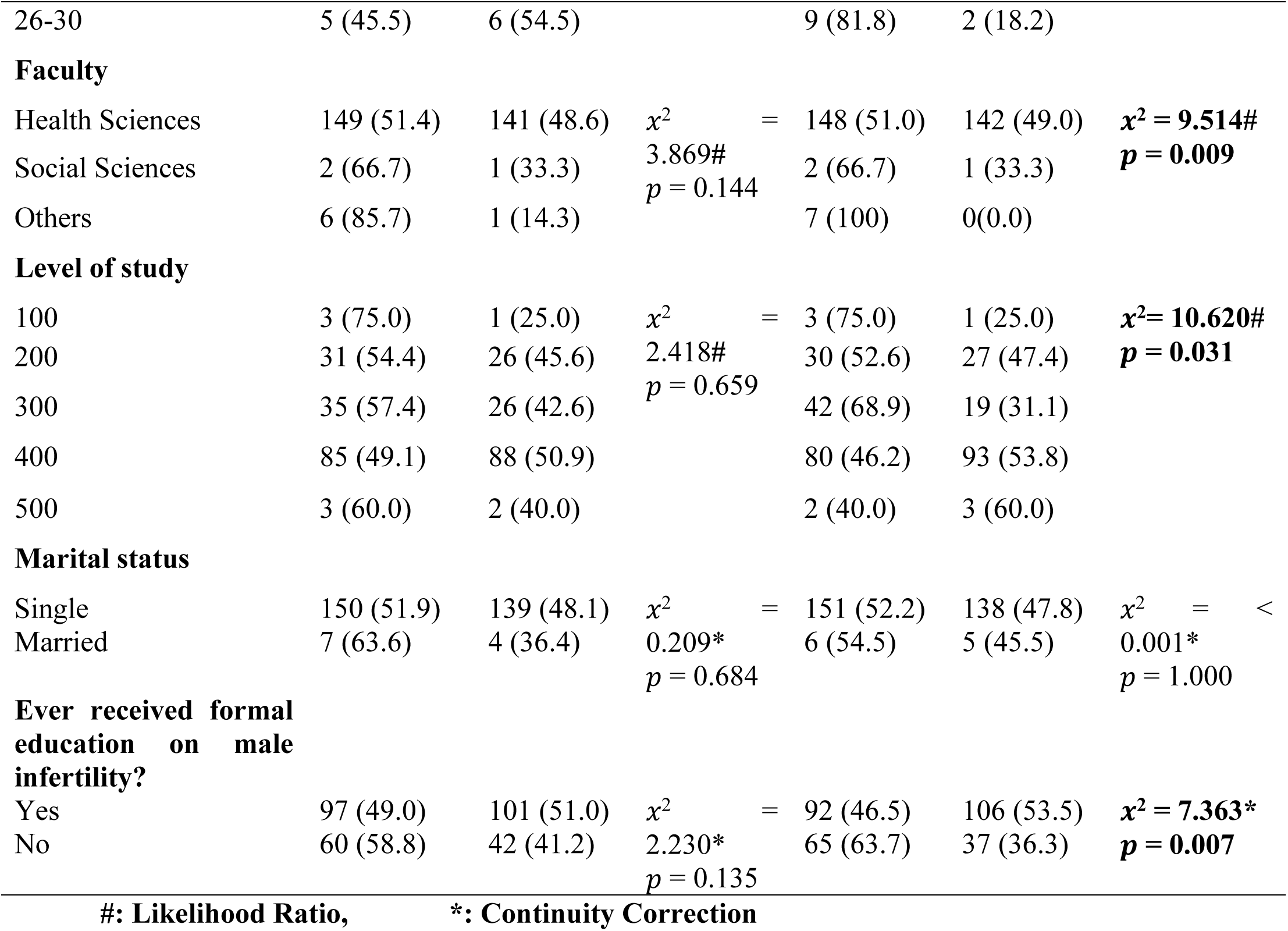
Association between the Respondents’ knowledge level of male infertility and their sociodemographic characteristics (N=300)

Regarding the attitude, Faculty affiliation showed a statistically significant association with attitude towards male infertility (p = 0.009), with Health Sciences respondents recording the highest proportion of positive attitudes (49.0%) compared to Social Sciences (33.3%) and other faculties (0.0%). Level of study was also significantly associated with attitude (p = 0.031); 400-level respondents had the most positive attitudes (53.8%), while 300-level respondents had the least (31.1%). Additionally, respondents who had received formal education on male infertility were significantly more likely to hold positive attitudes (53.5%) compared to those who had not (36.3%) (p = 0.007) (Table 3)

Table 4 above shows the binary logistic regression of the outcome variable ‘Knowledge level on male infertility’ and the selected socio-demographic predictors. None of the selected sociodemographic predictors was significant

**Table 4:** Binary Logistic Regression of the outcome variable ‘Knowledge of male infertility’ and the selected socio-demographic predictors.

| Variable | Sub-variable | Odds ratio | p-value | 95% confidence interval |  |
| --- | --- | --- | --- | --- | --- |
|  |  |  |  | Lower | Upper |
| **Gender** | Male | 1 |  |  |  |
|  | Female | 0.870 | 0.591 | 0.524 | 1.445 |
| **Age** | 16-20 | 1 |  |  |  |
|  | 21-25 | 1.150 | 0.586 | 0.696 | 1.900 |
|  | 26-30 | 1.528 | 0.527 | 0.411 | 5.675 |
| **Marital status** | Single | 1 |  |  |  |
|  | Married | 0.752 | 0.721 | 0.158 | 3.589 |
| **Ever received formal education on male infertility?** | Yes | 1 |  |  |  |
|  | No | 0.668 | 0.172 | 0.374 | 1.192 |

**Table 5:**
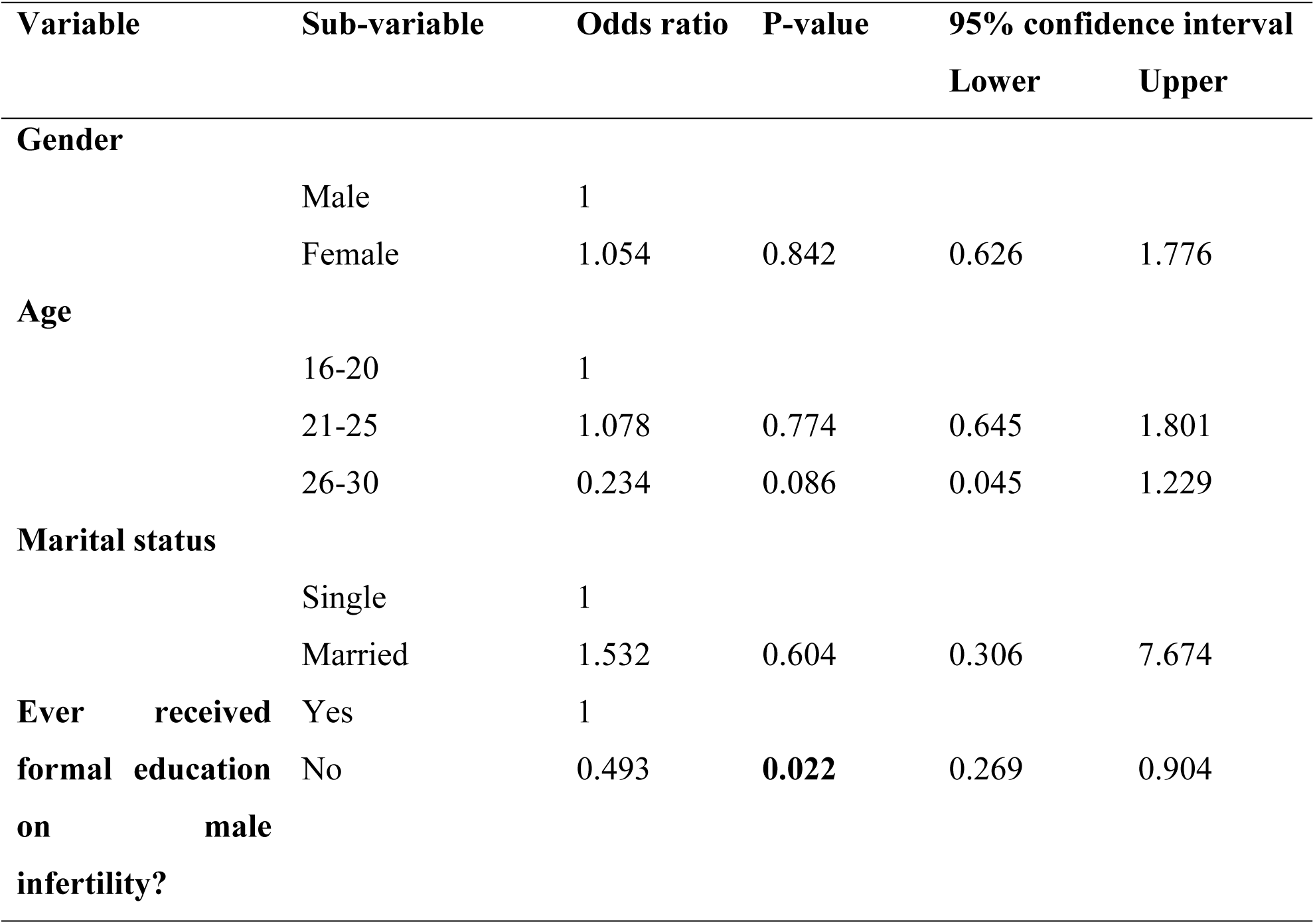
Binary Logistic Regression of the outcome variable ‘Attitude towards intimate partner violence’ and the selected socio-demographic predictors.

Table 4 above shows the binary logistic regression of the outcome variable ‘Attitude level on marital status’ and the selected socio-demographic predictors. For the variable ‘ever received formal education on male infertility, with “yes” as reference odds ratio 0.493, p= 0.022 (p<0.005), confidence interval, (0.269-0.904). This means that respondents that do not receive formal education on male infertility are two times less likely (1/0.493) or have 50.7% lower odds of having positive attitude towards male infertility compared to those who had received formal education.

### Qualitative Study

The qualitative reports employ a thematic analysis of a focus group discussion collected from undergraduates in Osun State University through coded interview transcripts. The analysis explores three core dimensions: knowledge, attitude, and perception regarding male infertility. The data reveal significant insights into how young adults understand, emotionally respond to, and socially contextualize male infertility in a Nigerian academic setting.

## KNOWLEDGE OF MALE INFERTILITY

### Definition and recognition of male infertility

Most participants recognized male infertility as a biological condition affecting men’s reproductive capacity. Definitions were often functional and intercourse-focused.

B1 defined it simply: “*inability of a man to impregnate a woman*.” B7 offered a more clinical description: “*the inability of a man to get a woman pregnant while having unprotected penetrative sex.”* B7 added a time-based criterion: “*the inability of a man to cause pregnancy after 12 months of regular unprotected penetrative vaginal sexual intercourse*.”

### Perceived causes of male infertility

Participants identified multiple causes, often blending medical, lifestyle, and cultural factors.

B2 listed: “*low sperm count, infection, lifestyle factors, genetic disorders, use of traditional medicine.”* B8 expanded on lifestyle influences: *“lifestyle factors like smoking, alcohol consumption, herbal consumption?”* (affirmed by respondents).

Infections, especially sexually transmitted infections, were consistently noted across files (b4, b7).

### Treatment possibilities and options

There was a strong consensus that male infertility is treatable, with awareness of both medical and assisted reproductive technologies.

B3, B4, B5, B6 all recorded unanimous “*yes*” responses to: “*Can male infertility be treated*?” B2 listed treatment modalities: “*medication, surgery, assisted reproductive techniques*.”

However, B7 and b8 highlighted scepticism toward traditional herbs, with respondents answering “*no*” to their efficacy.

## ATTITUDES TOWARD MALE INFERTILITY

Attitudes reflected societal biases, personal judgments, and emotional responses toward men with infertility.

### Infertility as a shared vs. Gendered issue

Participants strongly rejected the notion that infertility is primarily a female issue.

Across b1–b6, responses to “*Do you believe male infertility is more of a female issue than a male issue?”* Were a resounding “*noooo*.” B7 affirmed: *“no*” to the same question.

### Impact on masculinity and self-worth

Infertility was often seen as a threat to masculine identity, though opinions were divided.

B4 and b6 respondents said “*no*” to “*Do you think a man’s infertility reduces his masculinity?”* B5 and some in B1 and B3 said “*yes*.” B7 (respondent 5) offered a nuanced view: “*I think infertility is about bringing forth children; I won’t say children are the sign of masculinity.”* B7 (respondent 3) linked it to self-esteem: *“Also, due to the fact that the person is infertile, the person will have low self-esteem.”*

### Willingness to marry and social acceptance and medical help-seeking

A majority expressed reluctance to marry someone diagnosed with male infertility.

B1, b3, b5, b6, b7, b8 respondents answered “*no*” to “*Would you be willing to marry someone diagnosed with male infertility?”* B2 showed hesitation: “*not sure.*” B4 had mixed responses: “*no, yes.”* Despite stigma, there was strong support for men to seek medical help. B2, b3, b4, b5, b6, b7, b8 all recorded “*yes*” to “*Do you think men with infertility should seek medical help?”*

## PERCEPTION OF MALE INFERTILITY

### Male infertility as a taboo topic

Participants unanimously felt that male infertility is not widely discussed in Nigerian society. B1–b8 “*noooo*” to “Do you think male infertility is widely discussed in Nigerian society?”

### Major barriers to seeking treatment

Multiple barriers were identified, with stigma and cost being prominent.

B1 listed: “*stigma and cultural belief? Lack of awareness? High cost of treatment? Religious belief? Fear of being judged by family and friends?”* With respondents selecting “*all of the above.”* B3, b4, b5, b6 repeated similar barriers. B8 confirmed each barrier individually, including “*fear of being judged by families and friends?”* With a “*yes*.”

### Recommendations for improving awareness and access

Participants called for education, advocacy, and systemic support.

B2 recommended: “*health education, organisation of more awareness programs, public health education.”* B4, B5, B6 suggested “*establishment of more awareness, health education and check-ups/workshop.”* B7 advocated for outreaches: “*We should hold outreaches so that people will begin to know and understand that males are important as females when it comes to this issue.*” B7 proposed commemorative days: *“Although she has said what I wanted to say, but like we have world cancer day, world mental health day, I don’t think there is anything on male infertility?”* B7 (respondent 5) suggested institutional integration: “*In addition to what he said, court marriages can make it part of their procedural thing that both the wife and the husband should do a checkup, sperm count, pelvic ultrasound, before they proceed*.”

### Gender blame and social accountability

A subtle but recurring sub-theme was the tendency to blame women for infertility issues, even when acknowledging male factors. B7 (respondent 3) observed: “*Anytime there is infertility issue in the family, they tag the woman as a witch. Nobody even talks to the man as the problem.”* B7 (respondent 4) added: “*When there is a fertility problem, they always tell the man to marry another wife, but there needs to be awareness and education on the fact that infertility is both a male and female factor.”*

## DISCUSSION

This study assessed the knowledge, attitude, and perception of male infertility among undergraduates at Osun State University, Nigeria. The predominantly female sample (64.0%) reflects general enrollment trends in Nigerian health sciences [14] and is a common limitation in such studies [15]. A significant finding was the low overall knowledge (47.7%) and the predominance of negative attitudes (52.3%) towards male infertility, indicating a critical gap in reproductive health awareness. This is particularly concerning given the documented high burden of male infertility in Osogbo, where studies have reported that 84.3% of men attending fertility clinics have at least one abnormal seminal fluid parameter [6], and common abnormalities include asthenospermia (66.9%), teratospermia (63.9%), and oligospermia (21.0%) [11].

The functional understanding of male infertility demonstrated by participants, including its definition and causes, aligns with global clinical literature [1, 5]. Participants identified lifestyle factors such as smoking and alcohol consumption as causes, which is consistent with epidemiological findings identifying these as significant risk factors for male infertility (OR = 3.300 and OR = 4.990, respectively) [6]. The high awareness of treatment options, including ART, and the skepticism towards traditional remedies are encouraging signs of critical thinking, contrasting with findings from some other African studies [16]. However, the low overall knowledge score suggests this understanding is superficial and not comprehensive, highlighting the need for more structured educational interventions.

The attitudes expressed by students reveal a society in transition. The unanimous rejection of infertility as a “female-only” issue is a progressive shift from earlier studies that highlighted deep-seated gender biases [14]. This change may be attributed to the university environment and exposure to social media, which can challenge traditional beliefs [18]. However, the deep-seated cultural norms surrounding marriage and masculinity remain potent. The majority’s reluctance to marry an infertile man underscores the persistent social value placed on male fertility within patriarchal structures [15, 19]. This creates a painful paradox: men are no longer solely blamed, but they remain stigmatized and seen as less marriageable. This finding is particularly significant in light of qualitative evidence from fertility centers in Osogbo, which revealed that stigma often results in male infertility being perceived as a female issue, leading to delayed diagnosis and treatment [4]. The divided opinions on whether infertility diminishes masculinity reflect an ongoing internal negotiation of male identity in modern Nigerian society.

Perceptually, the silence surrounding male infertility was a dominant theme. The overwhelming agreement that it is not widely discussed confirms its status as a taboo, a silence that perpetuates misinformation and delays treatment-seeking [4]. The barriers identified—stigma, cost, and fear of judgment—are well-documented in low- and middle-income countries [15, 20]. The qualitative finding that women are still disproportionately blamed for infertility, despite intellectual acknowledgment of male factors, highlights the deep entrenchment of these cultural scripts. This “gender blame” underscores the urgent need for public education that frames infertility as a shared medical and social responsibility, particularly given the documented psychosocial consequences of male infertility, which include psychological distress, emotional turmoil, and social marginalization [4].

### Strength and limitation

The study’s strengths lie in its mixed-methods approach, which provided a nuanced understanding of the issue. The qualitative findings enriched the quantitative data by explaining the “why” behind the numbers. However, the findings must be considered in light of limitations. The sample was predominantly from the Faculty of Health Sciences, which may have positively biased knowledge levels and limits generalizability to students in other disciplines. Social desirability bias may have influenced responses on sensitive topics.

### Implication for study

Despite these limitations, the study has significant implications. The significant association between formal education and positive attitudes is a powerful testament to the potential of targeted educational interventions. It suggests that integrating comprehensive, gender-inclusive reproductive health education into university curricula, beyond just health sciences, is a critical step forward. Given the high prevalence of modifiable risk factors such as smoking and alcohol consumption among the general population, early educational interventions targeting young adults before they enter their peak reproductive years could have substantial preventive benefits. Participants’ own recommendations for awareness campaigns, workshops, and institutionalizing premarital screening provide a clear roadmap for future interventions that could help reduce the stigma and diagnostic delays documented in clinical settings.

## Conclusion

This study provides valuable insights into the knowledge, attitudes, and perceptions of male infertility among undergraduates at Osun State University. While students demonstrate a solid understanding of the biological and medical aspects of male infertility, their attitudes remain influenced by deep-seated cultural norms surrounding marriage, masculinity, and gender roles. Male infertility is perceived as a silenced issue, compounded by stigma, financial barriers, and societal blame directed disproportionately at women.

The findings highlight the critical need for integrated reproductive health education within university curricula, targeted awareness campaigns, and policy measures to reduce the cost and stigma associated with infertility care. By fostering open dialogue, promoting gender-inclusive narratives, and strengthening healthcare support systems, it is possible to transform perceptions and improve outcomes for individuals and couples affected by infertility in Nigeria and similar settings globally.

## Data Availability

All relevant data are within the manuscript

## Consent for publication

Not applicable

## Clinical trial

Not applicable

## Availability of data and material

The data for this study is provided within the manuscript

## Author’s contribution

KA, ATW and SCA worked on the study design; ATW, AOF, BTA, VAA and RF collected data; ODA, JF and EDO analyzed the data; KA and SCA supervised the project; EDO, ATW and SCA were major contributors in writing the manuscript. All authors read and approved the final manuscript.

## Acknowledgements

The authors greatly appreciate management of Osun State University and member of staff and students that contributed to the success of the study.

